# Air Pollution Increases Influenza Hospitalizations

**DOI:** 10.1101/2020.04.07.20057216

**Authors:** Gregor Singer, Joshua Graff Zivin, Matthew Neidell, Nicholas Sanders

## Abstract

Seasonal influenza is a recurring health burden shared widely across the globe. We study whether air quality affects the occurrence of severe influenza cases that require inpatient hospitalization. Using longitudinal information on local air quality and hospital admissions across the United States, we find that poor air quality increases the incidence of significant influenza hospital admissions. Effects diminish in years with greater influenza vaccine effectiveness. Apart from increasing vaccination rates, improving air quality may help reduce the spread and severity of influenza.

---

Seasonal influenza is a global health threat, with an average of 3-5 million severe cases per year and 290,000 to 650,000 respiratory deaths (*1, 2*). The disease exhibits variability in spread and severity across individuals, regions, and over time. Prior research has produced two broad sets of findings to explain this variation: a) meteorological factors that affect the spread of the virus, such as temperature, sunlight and humidity (*3, 4, 5, 6, 7, 8*); and b) individual level host factors, such as age, sex, underlying health and smoking that affect the intensity of symptoms (*9, 10*). We know considerably less, however, about how air pollution affects influenza spread and severity, a surprising gap given the pervasiveness of air pollution around the world and the well-established policy tools available to control it.

Air pollution could affect influenza hospitalizations via both susceptibility and exposure (*11*). Like smoking (*10*), air pollution can impair the respiratory functioning of patients, e.g., by damaging the respiratory epithelium, thereby facilitating the progression of influenza virus beyond the epithelial barrier into the lungs (*12, 13, 14, 15*). Existing medical research finds exposing *in vitro* respiratory epithelial cells to air pollution increases susceptibility and penetration of influenza (*13*), and experimental exposure of mice to air pollution before influenza infections increased morbidity and mortality (*16, 17*). Like humidity and temperature (*5, 6, 7, 18, 19*), air pollution particles could also impact the airborne survival of viruses outside the body (*18, 20, 21, 22, 23, 24*) and thus increase the probability of disease transmission.

We build on the existing evidence that links ambient air pollution with influenza spread and severity (*13, 16, 17, 21, 25, 26, 27, 28, 29, 30, 31, 32, 33, 34, 35*) with two significant advancements toward improving causal inference (*36*). First, we exploit a long panel of influenza-specific hospital admissions from numerous states across the United States (U.S.) to estimate statistical models that exploit both spatial and temporal variation within counties over time, limiting threats from confounding factors. Second, to better understand the causal link, we explore the role of the influenza vaccine in moderating this relationship. If the vaccine reduces infections and the probability of influenza spread, seasons in which the vaccine is more effective should weaken the link between air pollution and influenza (*37*).

Our analysis utilizes patient level data on inpatient hospitalization (*39*), which allows us to focus on severe cases specifically limited to influenza (for details on data, descriptives, and empirical methods see Supplementary Appendix S.1, S.2 and S.3). Our principal outcome of interest is the number of inpatient admissions per county-month where the primary diagnosis is influenza according to the International Classification of Diseases (ICD) (*40*). We combine this with high frequency air pollution readings of local ground monitors across the U.S., as well as data on local temperature, specific humidity, precipitation and wind speed (*41*). The richness of our data allows us to control for a wide variety of both regional and temporal controls. Our preferred specification includes county-by-year and month-by-year fixed effects. County-by-year effects control for differences in unobserved characteristics such as demographics, socio-economic factors, and health care access and protocols that influence pollution exposure and health outcomes across counties separately for each year. The month-by-year fixed effects control for general monthly and seasonal trends within each year in both influenza and pollution (*42*).

As our measure of pollution, we use the U.S. Environmental Protection Agency’s Air Quality Index (AQI), which we aggregate to county-by-month-by-year to match outcomes. The AQI is a measure of overall air quality based on the primary criteria pollutants specified in the Clean Air Act. Aggregation of pollutants means there are no real “units” for the measure. It is designed such that higher AQI values indicate worse air quality. To ensure we capture exposure to air pollution before diagnosis, we lag the AQI by one month. In all of our analyses, we focus on the influenza season (October to March). Figure 1 shows the seasonality of inpatient hospitalizations in our data (Figure 1a), which matches closely with general influenza-like illnesses reported by the Centers for Disease Control and Prevention (CDC) (Figure 1b). Figure 1c shows the age distribution of hospital admissions, which has important implications for vaccine effectiveness, described in more detail below.

**Figure 1:**
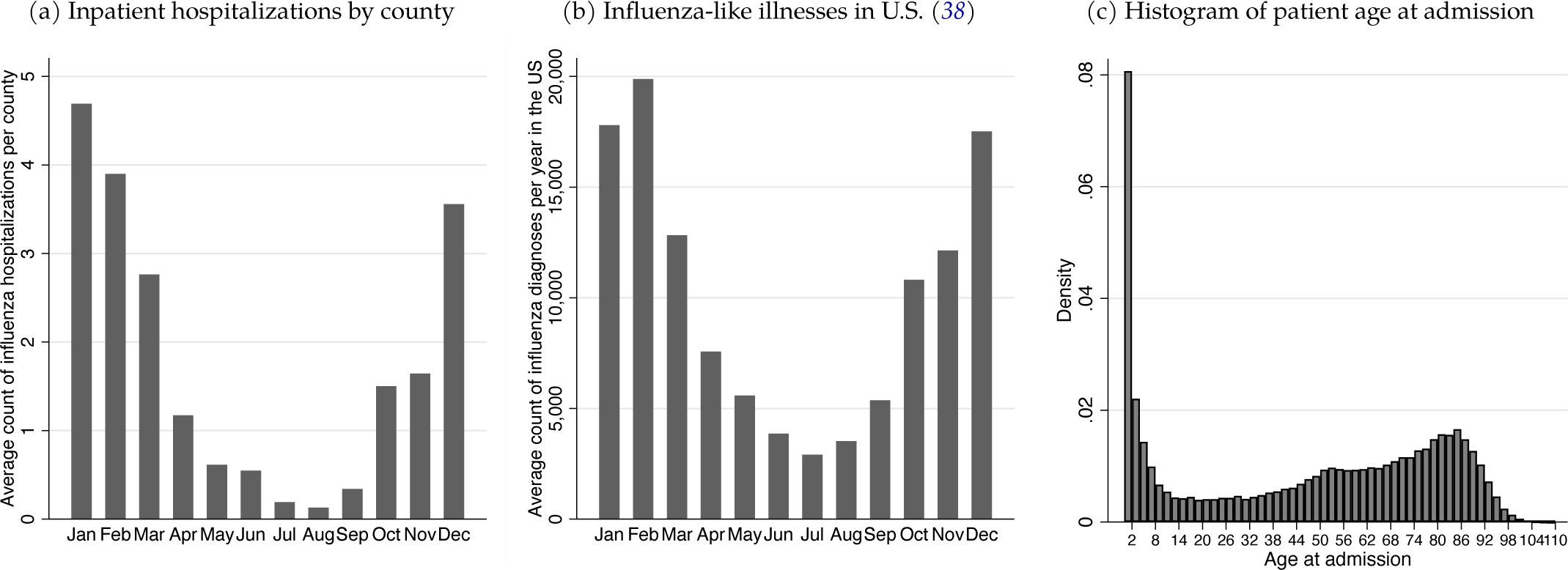
Influenza hospitalizations and influenza-like illnesses The left figure is based on the (*39*) inpatient data and shows the average count of influenza hospitalizations per county-month. The middle figure shows the distribution of recorded influenza-like illnesses from (*38*), which includes non-hospitalized cases. Data are pooled across the U.S. spanning 1997-2019. Not all health providers report to the Influenza-Like Illness (ILI) Network, and the number of providers reporting grew over time so total number of cases is a lower bound of true infection rates. The right figure shows the histogram of influenza inpatient admission ages pooling across states and time.

Figure 2a shows a clear positive correlation between air quality and count of influenza admissions in the raw data; higher AQI correlates with more influenza admissions (*43*). Figure 2b shows the correlation after adjusting both variables for fixed effects and weather controls. After this adjustment, a strong, positive correlation remains.

**Figure 2:**
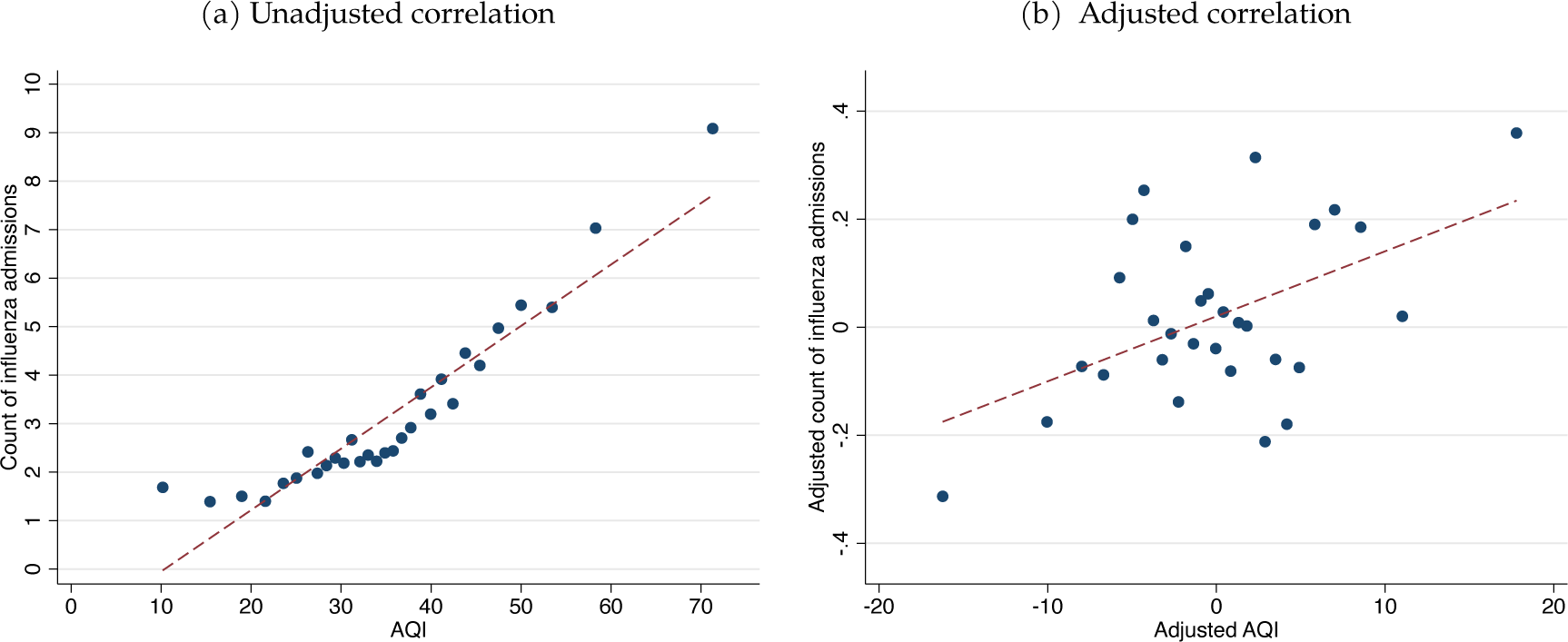
Influenza diagnosed hospital admissions and air pollution (AQI) The figures show binned scatterplots with 30 bins and a linear regression on the underlying hospitalization data of all age groups. The left figure shows the pooled raw data correlation. The right figure shows the correlation net of county-by-year and month fixed effects as well as weather controls.

Table 1 shows estimates from Poisson Pseudo-Maximum Likelihood regressions given the count nature of the dependent variable. The coefficients represent the change in the expected log of inpatient admission counts, which approximates a percentage change in number of county-year-month admissions within our data (*44*). Column (1) implies a 1-unit increase in the lagged monthly AQI results in a 0.56% increase in inpatient influenza admissions. To put this estimate in national context, a one standard deviation increase in AQI (12.79-unit increase in our data) amounts to approximately 4,064 additional inpatient hospitalizations for the 6-month influenza season in the U.S. (*45*).

**Table 1:** The effect of air pollution on hospitalizations

|  | Baseline effect |  | Interaction with vaccine effectiveness |  | Falsification outcome | Hospital charges |
| --- | --- | --- | --- | --- | --- | --- |
|  | (1) | (2) | (3) | (4) | (5) | (6) |
| AQI | 0.00560***<br>(0.002) |  | 0.0279**<br>(0.012) | 0.0138***<br>(0.005) | 0.0000921<br>(0.000) | 4929.4***<br>(1593.895) |
| Number of days AQI $\geq 100$ | | 0.0491**<br>(0.023) | | | | |
| AQI X Vaccine Effectiveness $\leq 8y$ | | | -0.0524**<br>(0.025) | | | |
| AQI X Vaccine Effectiveness $\geq 65y$ | | | | -0.0408**<br>(0.018) | | |
| Observations | 27448 | 27448 | 10699 | 10699 | 31307 | 12630 |
| Model | Poisson | Poisson | Poisson | Poisson | Poisson | OLS |
| Mean of outcome | 3.01 | 3.01 | 3.01 | 3.01 | 86.24 | 175613 |
| Mean of AQI predictor | 35.46 | 0.41 | 35.46 | 35.46 | 35.48 | 35.46 |
| Mean of vac. eff. | - | - | 0.47 | 0.28 | - | - |
| Weather controls | Yes | Yes | Yes | Yes | Yes | Yes |
| FE County X Year | Yes | Yes | No | No | Yes | Yes |
| FE Month | Yes | Yes | Yes | Yes | Yes | Yes |
| FE County X Inf. Seas. | No | No | Yes | Yes | No | No |

Notes: The dependent variable in Columns (1)-(4) is the count of hospital admissions with diagnosed influenza within a county-month. The dependent variable in Column (5) is the combined count of diabetes mellitus with complications, urinary tract infections, skull and face fractures, and osteoarthritis. The dependent variable in Column (6) is the dollar value (in 2018 US\$) of hospital charges for diagnosed influenza inpatients. We limit analysis to the influenza intensive months of October through March. Data measure vaccine effectiveness between 0 (low) and 1 (high) for the reported age groups, and are available from 2007 onward. The results are from a Poisson Pseudo-Maximum Likelihood or OLS regression, as indicated, with specified fixed effects and control variables. The number of included observations can vary across different outcomes due to fixed effects and varied counts in each county-month cell. Temperature controls consist of three separate bins, specific humidity controls consist of five separate bins, precipitation and wind speed are linear terms. All weather variables are based on county-month averages. The air quality index (AQI) is lagged one month. A higher AQI means worse air quality. We cluster standard errors (in parentheses) at the county level.

Column (2) replaces our continuous measure of air quality with the count of days in a month with air quality the EPA classifies as “unhealthy for sensitive groups” (AQI ≥ 100). These days are rare: in our data, the average county has around 0.4 such days per month. An additional unhealthy air quality day raises admission counts by approximately 5%. Continuing with our U.S.-wide calculation, an additional unhealthy air quality day in each county generates 2,786 additional inpatient hospitalizations per influenza season.

We next interact our air quality measure with a measure of influenza vaccine effectiveness. Every year, the CDC reports results from small-scale studies of that season’s influenza vaccine effectiveness rate by age group (see details in Supplementary Appendix S.1). Based on the histogram in Figure 1c, we use the vaccine effectiveness for the two age groups traditionally susceptible to health complications from influenza: children up to 8 and adults 65 and older. This group comprises 65% of inpatient hospitalization in our data. Figure 3 shows the regression-adjusted relationship between AQI and influenza admissions separately in seasons of low vaccine effectiveness and high vaccine effectiveness for the up to 8-year-old group and 65-year-and-older group, as determined by a median sample split (*46*). For both age groups, the relationship between air quality and admissions rates flattens and effectively disappears in years of high vaccine effectiveness. Columns (3) and (4) of Table 1 show a similar story using a more continuous measure of vaccine effectiveness. A vaccine effectiveness of 53% for the up to 8-year-old group or 34% for the 65-year-and-older group nullifies the link between air pollution and influenza hospitalizations (*47*).

**Figure 3:**
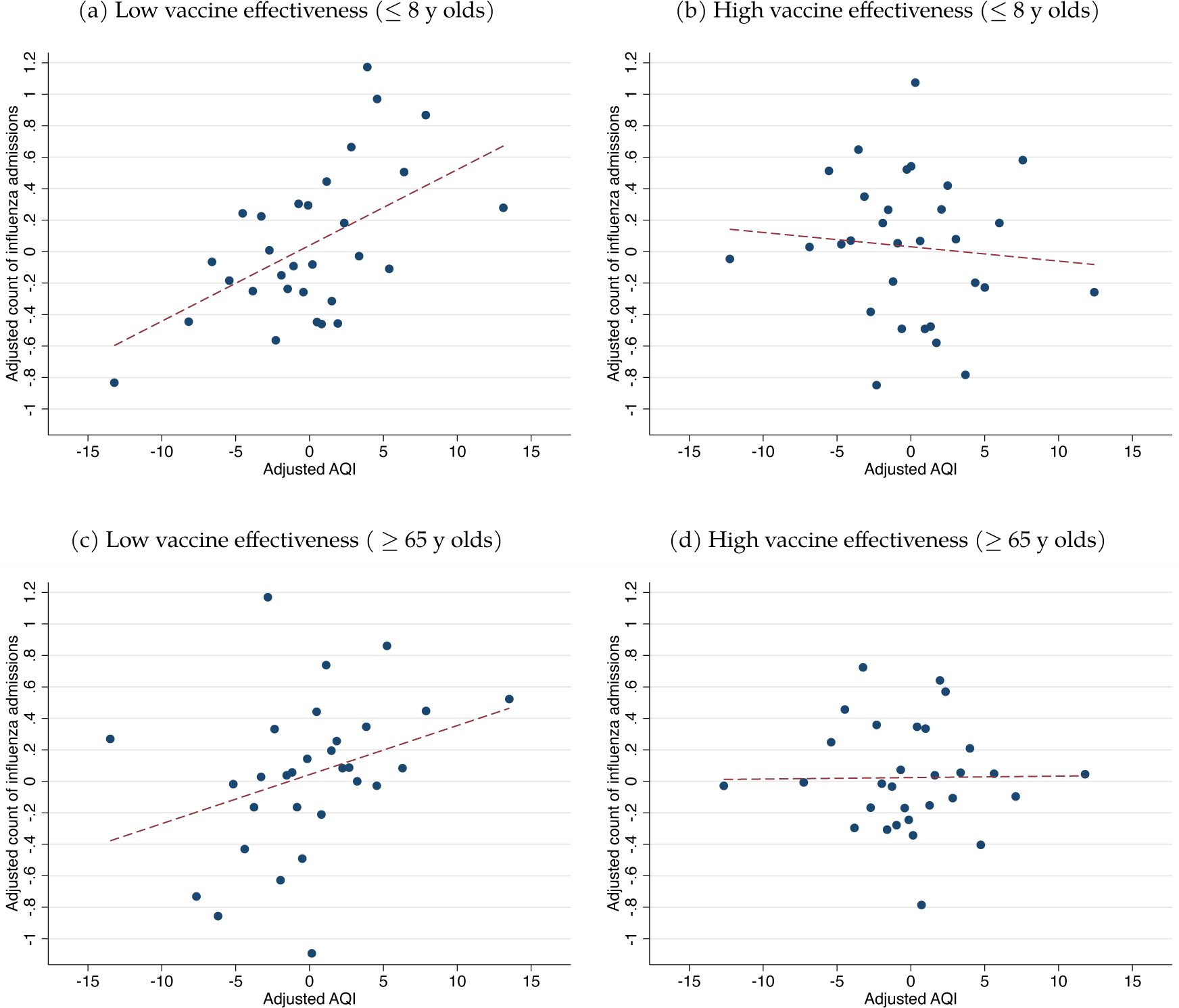
The effect of air pollution depends on vaccine effectiveness The figures shows binned scatterplots with 30 bins and a linear regression on the underlying data. Each shows the correlation net of county-by-year and month fixed effects as well as weather controls. Figures (a) and (b) show the relationship for seasons with below (a) and above (b) median vaccine effectiveness of the ≤ 8 year olds. Figures (c) and (d) show the same graphs for seasons with below (c) and above (d) median vaccine effectiveness for those 65 years and older.

While our fixed effects can address many unobservable factors, there remain possible confounders in establishing a causal link between pollution exposure and influenza hospitalizations. Air quality could trigger health problems in sensitive populations (e.g. asthmatics) who would then go to the hospital, where they might be observed to have influenza. For this reason, our analysis focused on patients whose *primary* diagnosis is influenza and ignore occurrences of influenza in secondary diagnoses. We also repeat our analysis using two alternative measures: patients where influenza is the *only* diagnosis and patients where *any* diagnosis is influenza. Supplementary Appendix S.4 shows that our results are robust to either of these alternatives.

We perform various falsification tests by repeating our analysis using health outcomes that should not correlate with air quality and health: diabetes mellitus with complications; urinary tract infections; skull and face fractures; and osteoarthritis (*48*). The result of a falsification test in Column (5), using the combined number of the above health outcomes, indicates a precise zero to the thousandth decimal place. We present estimates on each of these four falsification outcomes individually in Supplementary Appendix S.4 with similar results.

Supplementary Appendix S.4 explores heterogeneity and conducts further sensitivity analysis and robustness checks. Our estimates are stable across gender and age groups. We find suggestively larger effects for blacks and Hispanics, but the estimates are not statistically different from those for whites. We show robustness to (i) different weather controls, (ii) additional fixed effects, (iii) multilevel clustering of standard errors, (iv) different winsorization and interpolation of the raw AQI data, (v) including out-state patients at hospitals, (vi) focusing on states with a long time series only, (vii) using missing values instead of zeros for county-months with no hospital admissions, and (viii) using a linear ordinary least squares instead of a Poisson Pseudo-Maximum Likelihood estimator. We also show the effect of air pollution on outpatient hospitalization is larger than for inpatient hospitalizations, consistent with the notion that emergency department encounters are more frequent (but also less severe) than those requiring admission to the hospital.

As a final consideration, we shift from additional influenza cases to an economic endpoint. Column (6) of Table 1 shows ordinary least squares estimate of the effect of AQI on hospitalization charges for influenza admissions. This suggests a one-unit increase in AQI increases hospital billing by approximately $4,929 per month in the average county during influenza season. Across the U.S., a one standard deviation increase in AQI (12.79-unit increase) generates an additional $1.19 billion inpatient hospital charges per influenza season.

Using a rich longitudinal dataset, we provide causal evidence that air pollution increases hospitalization rates for seasonal influenza. Our findings offer novel evidence important for policy making, highlighting the heightened importance of increasing vaccination rates in polluted urban centers (*49*). This is especially important in developing countries, which house the most polluted cities in the world and have very low baseline vaccination rates (*50*). They also imply pollution controls can provide an important hedge against antigenic drift or shift in the influenza virus that renders the vaccine significantly less effective in some years, helping reduce global medical spending, avoid lost productivity, and reduce loss of human life.

If our results generalize to other respiratory viral infections, they will significantly understate the infectious-disease related benefits from environmental protection (*51*). They may also provide important insights for the ongoing fight against the COVID-19 pandemic (*52*). Social distancing and large scale reductions in economic activity aimed at reducing viral spread have also reduced air pollution (*53*), which may be helping reduce the impacts of the disease. As countries relax restrictions and economic activity resumes, they may choose to reduce environmental regulations in exchange for a more rapid return to economic growth: the U.S. EPA recently announced plans to suspend enforcement of environmental laws during the pandemic (*54*). Our results suggest there could be additional disease-related social costs to consider when worsening air quality during the economic recovery.

## Data Availability

The replication code and materials for both the manuscript and the supplementary materials will be made publicly available at Harvard Dataverse. The restricted access data can be accessed at https://www.hcup-us.ahrq.gov.

## ACKNOWLEDGEMENTS

We thank Luisa Osang and Jeffrey Shaman for helpful discussions. All errors are our own. The IRB for access to the HCUP data through the National Bureau of Economic Research (NBER) was approved by the NBER.

## Author contributions

GS, JGZ, MN and NS conceptualized the study, GS analyzed the data, and GS, JGZ, MN and NS wrote the manuscript.

## Data and materials availability

The replication code and materials for both the manuscript and the supplementary materials will be made publicly available at Harvard Dataverse. The restricted access data can be accessed at (*39*).

## SUPPLEMENTARY MATERIALS

## S.1 Data description

### Hospitalization data

We use hospital admission data from the Healthcare Cost and Utilization Project (HCUP) and focus on the inpatient data from hospital stays (*39*). We exploit patient level information on diagnosed diseases per International Classification of Diseases (ICD) codes, patient zip codes, admission months, age, gender, race as well as hospital charges. The data are available for a subset of U.S. states and years from 1991 (see Table S.1). We convert monetary hospital charges to common 2018 US$ using a GDP deflator from the World Bank (*60*).

To identify influenza hospitalizations, we count patients whose primary diagnosis is a strain of influenza. We use the Clinical Classifications Software (CCS) from the Agency for Healthcare Research and Quality (AHRQ) to classify relevant influenza ICD codes. These are the following ICD-9-CM codes: 4870, 4871, 4878, 488, 4880, 48801, 48802, 48809, 4881, 48811, 48812, 48819, 48881, 48882, 48889; and, for the period from October 2015 when the system was changed to ICD-10-CM, the following ICD-10-CM codes: J09X1, J09X2, J09X3, J09X9, J1000, J1001, J1008, J101, J102, J1081, J1082, J1083, J1089, J1100, J1108, J111, J112, J1181, J1182, J1183, J1189. We exclude patients whose primary diagnosis is not influenza, even if influenza is included among secondary diagnoses. Counting primary influenza diagnoses reflects a middle ground between two extreme alternatives for which we perform robustness checks. In one robustness check, we count patients who have any (primary or secondary) influenza diagnosis. In another robustness check we only count patients for whom influenza is their only diagnosis. We exclude patients whose zip code is from a different state than the hospital in which they are treated.

Hospitalization data are available at the patient zip code-by-month level, which we aggregate to the county-by-month level. We assign a zero value for admissions to counties in the months with no reported influenza admission. We only do this for counties and months in states that report data in the given year. During the influenza season from October to March, 57% of county-months have no influenza related hospital admissions in the HCUP data. Our results are robust with and without using the zero valued county-months in our estimations.

In four falsification tests, we use outcomes less likely to be affected by air quality: primary ICD codes associated with (i) diabetes mellitus with complications, (ii) urinary tract infections, (iii) skull and face fractures, and (iv) osteoarthritis. We use the categories and ICD codes from the Clinical Classifications Software (CCS) from the Agency for Healthcare Research and Quality (AHRQ). See Section S.1.1 for details. For a further robustness check, we use outpatient data from emergency departments (*61*) instead of the inpatient data, with the same strategy of counting influenza patients as above.

### Air quality

To measure air quality, we use the EPA Air Quality Index (AQI), which measures air quality derived from ground monitors (*62*). The AQI captures pollution from particulate matter (PM2.5), sulfur dioxide (SO2), carbon monoxide (CO), nitrogen dioxide (NO2) and ozone (O3). Further details on AQI calculation are provided by the EPA (*63*). We use the daily, county level, pre-aggregated data and further aggregate up to the county-by-month level. For missing county-months, we take the average value of the adjacent counties in the same month. We use the average value of the AQI within a month as well as the number of days with air at least “unhealthy for sensitive groups” according to the EPA (AQI≥100). We winsorize the AQI at the top and bottom 1% for the main analysis and show robust results without winsorization. For our analysis, we take the one month lagged AQI to identify exposure to air pollution before influenza diagnosis and not afterwards.

### Weather controls

We use pre-aggregated monthly weather averages from (*64, 65*), including temperature, specific humidity, wind speed, and precipitation, and aggregate grid points up to the county-by-month level.

### Influenza seasons

We use data on the timing of national influenza-like illnesses from the CDC (*38*) to identify the main influenza months: October through March (see Figure 1b). This coincides with the reported influenza season in various CDC publications. We restrict our main analysis to this influenza season.

### Vaccine effectiveness

We use the estimated vaccine effectiveness, for different age groups, by influenza season, from the CDC (*66*). Underlying cited studies are available from 2007/2008. Since vaccine effectiveness can vary across age groups during the same influenza season, we use the reported effectiveness of the two age groups most relevant for our study: children up to 8 years old and for people 65 years and older. Figure 1c shows these are the main age groups observed in the HCUP inpatient data with primary influenza diagnoses.

#### S.1.1 ICD codes for falsification tests

We use the categories from the Clinical Classifications Software (CCS) from the Agency for Healthcare Research and Quality (AHRQ) to identify the relevant ICD codes.

##### Diabetes mellitus with complications ICD-9-CM codes

24901, 24910, 24911, 24920, 24921, 24930, 24931, 24940, 24941, 24950, 24951, 24960, 24961, 24970, 24971, 24980, 24981, 24990, 24991, 25002, 25003, 25010, 25011, 25012, 25013, 25020, 25021, 25022, 25023, 25030, 25031, 25032, 25033, 25040, 25041, 25042, 25043, 25050, 25051, 25052, 25053, 25060, 25061, 25062, 25063, 25070, 25071, 25072, 25073, 25080, 25081, 25082, 25083, 25090, 25091, 25092, 25093;

##### Diabetes mellitus with complications ICD-10-CM codes

E0800, E0801, E0810, E0811, E0821, E0822, E0829, E08311, E08319, E08321, E083211, E083212, E083213, E083219, E08329, E083291, E083292, E083293, E083299, E08331, E083311, E083312, E083313, E083319, E08339, E083391, E083392, E083393, E083399, E08341, E083411, E083412, E083413, E083419, E08349, E083491, E083492, E083493, E083499, E08351, E083511, E083512, E083513, E083519, E083521, E083522, E083523, E083529, E083531, E083532, E083533, E083539, E083541, E083542, E083543, E083549, E083551, E083552, E083553, E083559, E08359, E083591, E083592, E083593, E083599, E0836, E0837X1, E0837X2, E0837X3, E0837X9, E0839, E0840, E0841, E0842, E0843, E0844, E0849, E0851, E0852, E0859, E08610, E08618, E08620, E08621, E08622, E08628, E08630, E08638, E08641, E08649, E0865, E0869, E088, E0900, E0901, E0910, E0911, E0921, E0922, E0929, E09311, E09319, E09321, E093211, E093212, E093213, E093219, E09329, E093291, E093292, E093293, E093299, E09331, E093311, E093312, E093313, E093319, E09339, E093391, E093392, E093393, E093399, E09341, E093411, E093412, E093413, E093419, E09349, E093491, E093492, E093493, E093499, E09351, E093511, E093512, E093513, E093519, E093521, E093522, E093523, E093529, E093531, E093532, E093533, E093539, E093541, E093542, E093543, E093549, E093551, E093552, E093553, E093559, E09359, E093591, E093592, E093593, E093599, E0936, E0937X1, E0937X2, E0937X3, E0937X9, E0939, E0940, E0941, E0942, E0943, E0944, E0949, E0951, E0952, E0959, E09610, E09618, E09620, E09621, E09622, E09628, E09630, E09638, E09641, E09649, E0965, E0969, E098, E1010, E1011, E1021, E1022, E1029, E10311, E10319, E10321, E103211, E103212, E103213, E103219, E10329, E103291, E103292, E103293, E103299, E10331, E103311, E103312, E103313, E103319, E10339, E103391, E103392, E103393, E103399, E10341, E103411, E103412, E103413, E103419, E10349, E103491, E103492, E103493, E103499, E10351, E103511, E103512, E103513, E103519, E103521, E103522, E103523, E103529, E103531, E103532, E103533, E103539, E103541, E103542, E103543, E103549, E103551, E103552, E103553, E103559, E10359, E103591, E103592, E103593, E103599, E1036, E1037X1, E1037X2, E1037X3, E1037X9, E1039, E1040, E1041, E1042, E1043, E1044, E1049, E1051, E1052, E1059, E10610, E10618, E10620, E10621, E10622, E10628, E10630, E10638, E10641, E10649, E1065, E1069, E108, E1100, E1101, E1110, E1111, E1121, E1122, E1129, E11311, E11319, E11321, E113211, E113212, E113213, E113219, E11329, E113291, E113292, E113293, E113299, E11331, E113311, E113312, E113313, E113319, E11339, E113391, E113392, E113393, E113399, E11341, E113411, E113412, E113413, E113419, E11349, E113491, E113492, E113493, E113499, E11351, E113511, E113512, E113513, E113519, E113521, E113522, E113523, E113529, E113531, E113532, E113533, E113539, E113541, E113542, E113543, E113549, E113551, E113552, E113553, E113559, E11359, E113591, E113592, E113593, E113599, E1136, E1137X1, E1137X2, E1137X3, E1137X9, E1139, E1140, E1141, E1142, E1143, E1144, E1149, E1151, E1152, E1159, E11610, E11618, E11620, E11621, E11622, E11628, E11630, E11638, E11641, E11649, E1165, E1169, E118, E1300, E1301, E1310, E1311, E1321, E1322, E1329, E13311, E13319, E13321, E133211, E133212, E133213, E133219, E13329, E133291, E133292, E133293, E133299, E13331, E133311, E133312, E133313, E133319, E13339, E133391, E133392, E133393, E133399, E13341, E133411, E133412, E133413, E133419, E13349, E133491, E133492, E133493, E133499, E13351, E133511, E133512, E133513, E133519, E133521, E133522, E133523, E133529, E133531, E133532, E133533, E133539, E133541, E133542, E133543, E133549, E133551, E133552, E133553, E133559, E13359, E133591, E133592, E133593, E133599, E1336, E1337X1, E1337X2, E1337X3, E1337X9, E1339, E1340, E1341, E1342, E1343, E1344, E1349, E1351, E1352, E1359, E13610, E13618, E13620, E13621, E13622, E13628, E13630, E13638, E13641, E13649, E1365, E1369, E138.

##### Urinary tract infections ICD-9-CM codes

03284, 59000, 59001, 59010, 59011, 5902, 5903, 59080, 59081, 5909, 5950, 5951, 5952, 5953, 5954, 59581, 59582, 59589, 5959, 5970, 59780, 59781, 59789, 59800, 59801, 5990;

##### Urinary tract infections ICD-10-CM codes

A0225, A3685, B3741, B3749, N10, N110, N111, N118, N119, N12, N136, N151, N3000, N3001, N3010, N3011, N3020, N3021, N3030, N3031, N3040, N3041, N3080, N3081, N3090, N3091, N340, N341, N342, N343, N390.

##### Skull and face fractures ICD-9-CM codes

80000, 80001, 80002, 80003, 80004, 80005, 80006, 80009, 80050, 80051, 80052, 80053, 80054, 80055, 80056, 80059, 80100, 80101, 80102, 80103, 80104, 80105, 80106, 80109, 80150, 80151, 80152, 80153, 80154, 80155, 80156, 80159, 8020, 8021, 80220, 80221, 80222, 80223, 80224, 80225, 80226, 80227, 80228, 80229, 80230, 80231, 80232, 80233, 80234, 80235, 80236, 80237, 80238, 80239, 8024, 8025, 8026, 8027, 8028, 8029, 80300, 80301, 80302, 80303, 80304, 80305, 80306, 80309, 80350, 80351, 80352, 80353, 80354, 80355, 80356, 80359, 80400, 80401, 80402, 80403, 80404, 80405, 80406, 80409, 80450, 80451, 80452, 80453, 80454, 80455, 80456, 80459, 9050;

##### Skull and face fractures ICD-10-CM codes

S020XXA, S020XXB, S020XXD, S020XXG, S020XXK, S020XXS, S02101A, S02101B, S02101D, S02101G, S02101K, S02101S, S02102A, S02102B, S02102D, S02102G, S02102K, S02102S, S02109A, S02109B, S02109D, S02109G, S02109K, S02109S, S0210XA, S0210XB, S0210XD, S0210XG, S0210XK, S0210XS, S02110A, S02110B, S02110D, S02110G, S02110K, S02110S, S02111A, S02111B, S02111D, S02111G, S02111K, S02111S, S02112A, S02112B, S02112D, S02112G, S02112K, S02112S, S02113A, S02113B, S02113D, S02113G, S02113K, S02113S, S02118A, S02118B, S02118D, S02118G, S02118K, S02118S, S02119A, S02119B, S02119D, S02119G, S02119K, S02119S, S0211AA, S0211AB, S0211AD, S0211AG, S0211AK, S0211AS, S0211BA, S0211BB, S0211BD, S0211BG, S0211BK, S0211BS, S0211CA, S0211CB, S0211CD, S0211CG, S0211CK, S0211CS, S0211DA, S0211DB, S0211DD, S0211DG, S0211DK, S0211DS, S0211EA, S0211EB, S0211ED, S0211EG, S0211EK, S0211ES, S0211FA, S0211FB, S0211FD, S0211FG, S0211FK, S0211FS, S0211GA, S0211GB, S0211GD, S0211GG, S0211GK, S0211GS, S0211HA, S0211HB, S0211HD, S0211HG, S0211HK, S0211HS, S02121A, S02121B, S02121D, S02121G, S02121K, S02121S, S02122A, S02122B, S02122D, S02122G, S02122K, S02122S, S02129A, S02129B, S02129D, S02129G, S02129K, S02129S, S0219XA, S0219XB, S0219XD, S0219XG, S0219XK, S0219XS, S022XXA, S022XXB, S022XXD, S022XXG, S022XXK, S022XXS, S0230XA, S0230XB, S0230XD, S0230XG, S0230XK, S0230XS, S0231XA, S0231XB, S0231XD, S0231XG, S0231XK, S0231XS, S0232XA, S0232XB, S0232XD, S0232XG, S0232XK, S0232XS, S023XXA, S023XXB, S023XXD, S023XXG, S023XXK, S023XXS, S02400A, S02400B, S02400D, S02400G, S02400K, S02400S, S02401A, S02401B, S02401D, S02401G, S02401K, S02401S, S02402A, S02402B, S02402D, S02402G, S02402K, S02402S, S0240AA, S0240AB, S0240AD, S0240AG, S0240AK, S0240AS, S0240BA, S0240BB, S0240BD, S0240BG, S0240BK, S0240BS, S0240CA, S0240CB, S0240CD, S0240CG, S0240CK, S0240CS, S0240DA, S0240DB, S0240DD, S0240DG, S0240DK, S0240DS, S0240EA, S0240EB, S0240ED, S0240EG, S0240EK, S0240ES, S0240FA, S0240FB, S0240FD, S0240FG, S0240FK, S0240FS, S02411A, S02411B, S02411D, S02411G, S02411K, S02411S, S02412A, S02412B, S02412D, S02412G, S02412K, S02412S, S02413A, S02413B, S02413D, S02413G, S02413K, S02413S, S0242XA, S0242XB, S0242XD, S0242XG, S0242XK, S0242XS, S025XXA, S025XXB, S025XXD, S025XXG, S025XXK, S025XXS, S02600A, S02600B, S02600D, S02600G, S02600K, S02600S, S02601A, S02601B, S02601D, S02601G, S02601K, S02601S, S02602A, S02602B, S02602D, S02602G, S02602K, S02602S, S02609A, S02609B, S02609D, S02609G, S02609K, S02609S, S02610A, S02610B, S02610D, S02610G, S02610K, S02610S, S02611A, S02611B, S02611D, S02611G, S02611K, S02611S, S02612A, S02612B, S02612D, S02612G, S02612K, S02612S, S0261XA, S0261XB, S0261XD, S0261XG, S0261XK, S0261XS, S02620A, S02620B, S02620D, S02620G, S02620K, S02620S, S02621A, S02621B, S02621D, S02621G, S02621K, S02621S, S02622A, S02622B, S02622D, S02622G, S02622K, S02622S, S0262XA, S0262XB, S0262XD, S0262XG, S0262XK, S0262XS, S02630A, S02630B, S02630D, S02630G, S02630K, S02630S, S02631A, S02631B, S02631D, S02631G, S02631K, S02631S, S02632A, S02632B, S02632D, S02632G, S02632K, S02632S, S0263XA, S0263XB, S0263XD, S0263XG, S0263XK, S0263XS, S02640A, S02640B, S02640D, S02640G, S02640K, S02640S, S02641A, S02641B, S02641D, S02641G, S02641K, S02641S, S02642A, S02642B, S02642D, S02642G, S02642K, S02642S, S0264XA, S0264XB, S0264XD, S0264XG, S0264XK, S0264XS, S02650A, S02650B, S02650D, S02650G, S02650K, S02650S, S02651A, S02651B, S02651D, S02651G, S02651K, S02651S, S02652A, S02652B, S02652D, S02652G, S02652K, S02652S, S0265XA, S0265XB, S0265XD, S0265XG, S0265XK, S0265XS, S0266XA, S0266XB, S0266XD, S0266XG, S0266XK, S0266XS, S02670A, S02670B, S02670D, S02670G, S02670K, S02670S, S02671A, S02671B, S02671D, S02671G, S02671K, S02671S, S02672A, S02672B, S02672D, S02672G, S02672K, S02672S, S0267XA, S0267XB, S0267XD, S0267XG, S0267XK, S0267XS, S0269XA, S0269XB, S0269XD, S0269XG, S0269XK, S0269XS, S0280XA, S0280XB, S0280XD, S0280XG, S0280XK, S0280XS, S0281XA, S0281XB, S0281XD, S0281XG, S0281XK, S0281XS, S0282XA, S0282XB, S0282XD, S0282XG, S0282XK, S0282XS, S02831A, S02831B, S02831D, S02831G, S02831K, S02831S, S02832A, S02832B, S02832D, S02832G, S02832K, S02832S, S02839A, S02839B, S02839D, S02839G, S02839K, S02839S, S02841A, S02841B, S02841D, S02841G, S02841K, S02841S, S02842A, S02842B, S02842D, S02842G, S02842K, S02842S, S02849A, S02849B, S02849D, S02849G, S02849K, S02849S, S0285XA, S0285XB, S0285XD, S0285XG, S0285XK, S0285XS, S028XXA, S028XXB, S028XXD, S028XXG, S028XXK, S028XXS, S0291XA, S0291XB, S0291XD, S0291XG, S0291XK, S0291XS, S0292XA, S0292XB, S0292XD, S0292XG, S0292XK, S0292XS.

##### Osteoarthritis ICD-9-CM codes

71500, 71504, 71509, 71510, 71511, 71512, 71513, 71514, 71515, 71516, 71517, 71518, 71520, 71521, 71522, 71523, 71524, 71525, 71526, 71527, 71528, 71530, 71531, 71532, 71533, 71534, 71535, 71536, 71537, 71538, 71580, 71589, 71590, 71591, 71592, 71593, 71594, 71595, 71596, 71597, 71598, V134;

##### Osteoarthritis ICD-10-CM codes

M150, M151, M152, M153, M154, M158, M159, M160, M1610, M1611, M1612, M162, M1630, M1631, M1632, M164, M1650, M1651, M1652, M166, M167, M169, M170, M1710, M1711, M1712, M172, M1730, M1731, M1732, M174, M175, M179, M180, M1810, M1811, M1812, M182, M1830, M1831, M1832, M184, M1850, M1851, M1852, M189, M19011, M19012, M19019, M19021, M19022, M19029, M19031, M19032, M19039, M19041, M19042, M19049, M19071, M19072, M19079, M19111, M19112, M19119, M19121, M19122, M19129, M19131, M19132, M19139, M19141, M19142, M19149, M19171, M19172, M19179, M19211, M19212, M19219, M19221, M19222, M19229, M19231, M19232, M19239, M19241, M19242, M19249, M19271, M19272, M19279, M1990, M1991, M1992, M1993.

## S.2 Additional descriptive statistics

Table S.1 contains states and years with available admission months and patient zip codes in the inpatient hospitalization data we use. Table S.2 contains summary statistics for inpatient hospital admissions with a primary influenza diagnosis, average monthly AQI per county-month, and the number of days with AQI ≥ 100. We use the standard deviation of the AQI during the influenza season (12.79) as well as the average inpatient hospitalization numbers (3.01) for the calculation of absolute effects based on our Poisson Pseudo-Maximum Likelihood estimation.

**Table S.1:** Data coverage with available zip codes and admission months

|  |  |
| --- | --- |
| Arizona | 1991,1992,1993,1994,1995,1996,1997,1998,1999,2000,2001,2002,2003,2004,2005,2006,2007,2008,2009,2010,2011,2012,2013,2014,2015,2016 |
| Arkansas | 2009 |
| Colorado | 2003,2004,2005,2006,2007,2009,2012 |
| Hawaii | 2009 |
| Iowa | 1999,2009 |
| Kentucky | 2005,2006,2007,2008,2009,2010,2011,2012,2013,2014 |
| Maryland | 2009,2010,2011,2012 |
| Massachusetts | 2007,2008,2009,2010,2011,2012,2013,2014 |
| Michigan | 2009 |
| New Jersey | 1991,1992,1993,1994,1995,1996,1997,1998,1999,2000,2001,2002,2003,2004,2005,2006,2007,2008,2009,2010,2011,2012,2013,2014,2015 |
| New York | 1993,1994,1995,1996,1997,1998,1999,2000,2001,2002,2003,2004,2005,2006,2007,2008,2009,2010,2011,2012,2013,2014,2015 |
| Oregon | 1999,2008,2009 |
| South Dakota | 2009 |
| Utah | 2009 |
| Vermont | 2009 |
| Washington | 1993,1994,1995,1996,1997,1998,1999,2000,2001,2002,2003,2004,2005,2006,2007,2008,2009,2010,2011,2012,2013 |
| Wisconsin | 1999,2009 |

Notes: The table shows the states and years used in the main analysis.

**Table S.2:**
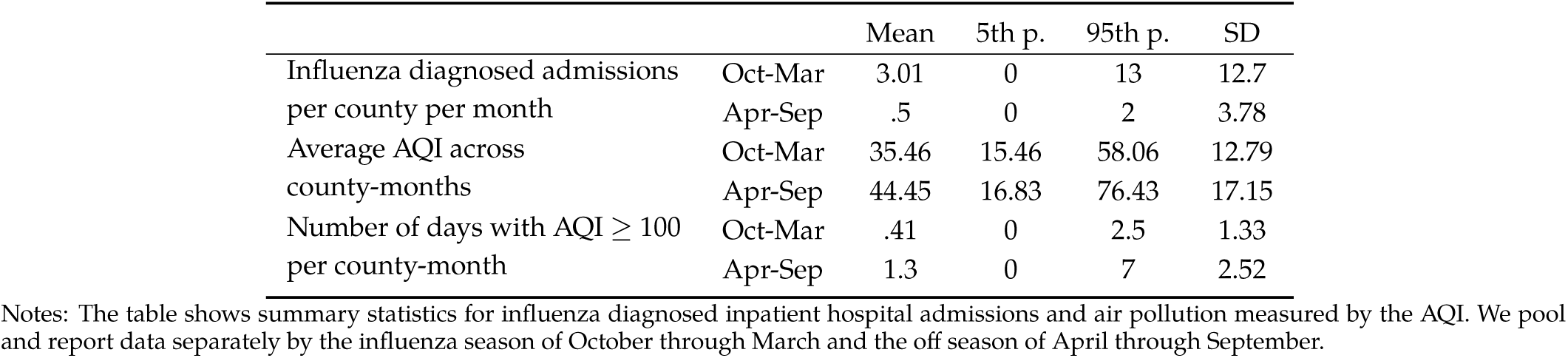
Summary statistics of influenza hospitalizations and air pollution (AQI)

## S.3 Empirical strategy

We estimate the relationship between influenza-related inpatient hospitalizations *H*_*cym*_ and the lagged air quality index *AQI*_*cym*−_1 at the county *c* by calendar month *m* by year *y* level using a Poisson model:

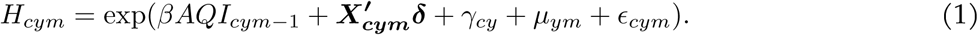

We include county-by-year fixed effects *γ*_*cy*_ to control for changing factors such as population size, income, demography and influenza testing procedures across counties and time. This also captures unobserved annual shocks at the county level that affect both air pollution and hospitalizations. Calendar month-by-year fixed effects *µ*_*ym*_ control for a flexible overall time trend. Results are robust to including additional fixed effects such as state-by-calendar month or county-by-influenza season fixed effects.

While county-by-year fixed effects capture the bulk of climatic differences across counties, we also control for within-year differences with a vector of weather control variables ***X***_***cym***_. This includes temperature, specific humidity, precipitation, and wind speed in various combinations. Temperature and humidity has been shown to affect both virus survival (see (*5, 6, 7, 19, 67*) and air pollution (*18, 23, 55*). In our baseline we include three temperature (C) bins (< 0, ≥ 0 & < 15 and > 15), five bins based on the quintiles of specific humidity, and linear terms for precipitation and wind speed.

We lag the AQI by one month to account for hospital admissions data at the monthly level. The goal is to capture pollution exposure prior to the influenza diagnosis, not after. In principle, air pollution could also affect patient progression after diagnosis, but we focus on the effect of pollution leading up to the diagnosis.

We estimate the model with a pseudo-maximum likelihood estimator (*68, 69*), which performs well with a large number of zeros and is consistent with over- or under-dispersion in the data (*70*). We cluster standard errors at the county level to allow for arbitrary heteroskedasticity and serial correlation in the errors, and show robustness to two-way clustering at the added state-year level.

## S.4 Additional results and robustness checks

Table S.3 provides falsification tests with outcomes unlikely to be correlated with air pollution. Column 1 repeats our baseline results for influenza patients. The next four columns use inpatient hospitalizations with a primary diagnosis of diabetes mellitus with complications, urinary tract infections, skull and face fractures, and osteoarthritis. Coefficients and standard errors indicate a precise zero effect for these outcomes.

Table S.4 explores heterogeneous effects by age, gender and race. Estimates across different groups are statistically indistinguishable from one another, however, the point estimates for blacks and especially Hispanics are larger than for whites.

Table S.5 explores robustness of our main results to different controls, fixed effects, and standard error calculations. Column (1) replicates the baseline results, and reports the estimates for our weather controls (reporting was suppressed in the manuscript for simplicity). Temperature and humidity controls are included as dummies for separate bins. While the coefficients on temperature are not statistically significant (county-year fixed effects absorb much of the large-scale variation), the sign is as expected. Temperatures below zero C as well as above 15 C lead to fewer observed hospitalizations (see also (*5, 6*)). Humidity decreases hospitalizations consistent with (*5, 6, 7, 19*), while precipitation and wind speed have no statistically or economically significant effects.

In Column (2) of Table S.5, we drop weather controls, and in Column (3) we include alternative functional forms of the weather controls using second order polynomials in temperature and humidity with a full set of interactions. In Columns (4) and (5) we include county-by-influenza season (Oct - Mar) fixed effects and state-by-month of the year fixed effects. Columns (6) and (7) replicate (1) and (4), but cluster standard errors on the county level as well as on the state-by-year level to allow additional arbitrary spatial correlation of errors across counties within a state-year.

Table S.6 reports from further robustness checks. Column (1) replicates the baseline results. Column (2) does not winsorize the AQI data. Column (3) drops county-month cells with missing AQI measures (rather than interpolating them based on the average value of the adjacent counties). Column (4) includes patients whose zip code is from a different state than the hospital in which they are treated. Column (5) restricts to states with at least seven years of reported data: Arizona, Colorado, Kentucky, Massachusetts, New Jersey, New York, and Washington. Column (6) drops county-months with no reported influenza admissions (rather than assigning a zero value for admissions). Column (7) contains results from an ordinarily least square (OLS) regression instead of a Poisson Pseudo-Maximum Likelihood regression.

Columns (8) and (9) use alternative assumptions on who to count as an influenza patient. Our baseline only counts patients whose primary diagnosis is influenza. Column (8) counts patients where all diagnoses are influenza, i.e., there are no other diagnosed conditions. Column (9) counts all patients with any influenza diagnosis, primary or non-primary. Columns (10) and (11) use the data on outpatient (instead of inpatient) hospitalizations as the outcome variable. The effect of AQI is slightly larger on outpatient hospitalizations, consistent with the notion that these are less severe but more frequent than inpatient hospitalizations.

**Table S.3:**
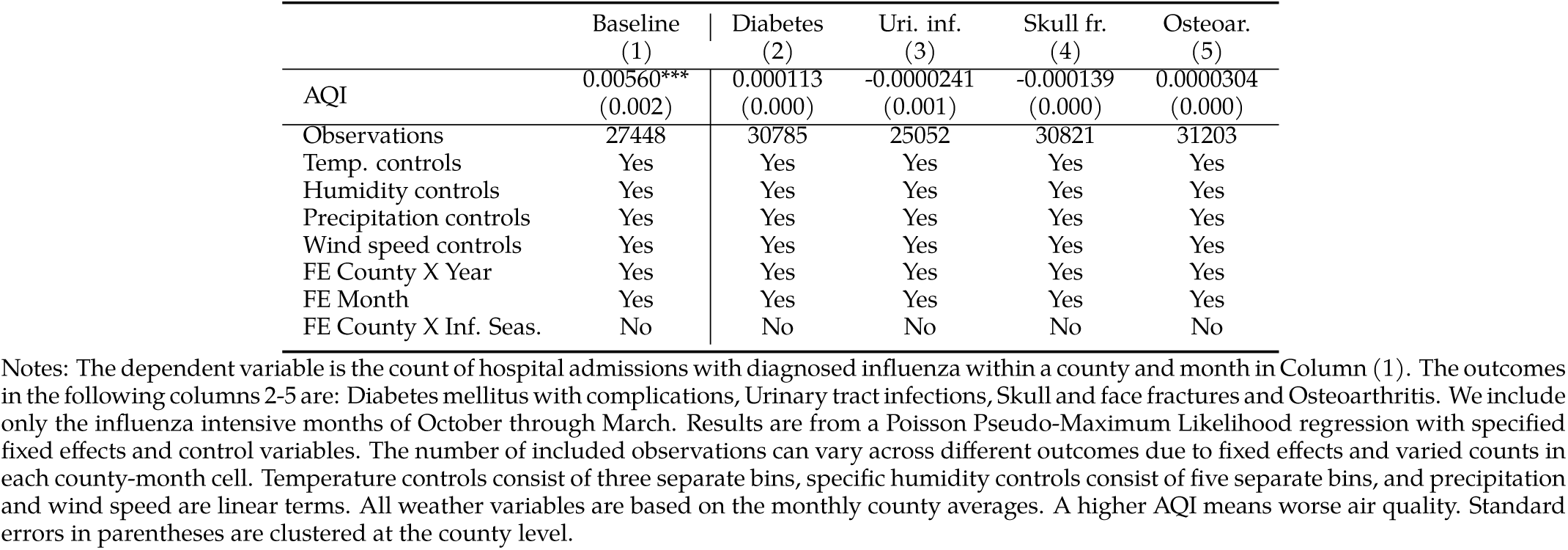
The effect of air pollution on non-related hospitalizations:

|  | Baseline<br>(1) | Diabetes<br>(2) | Uri. inf.<br>(3) | Skull fr.<br>(4) | Osteoar.<br>(5) |
| --- | --- | --- | --- | --- | --- |
| AQI | 0.00560***<br>(0.002) | 0.000113<br>(0.000) | -0.0000241<br>(0.001) | -0.000139<br>(0.000) | 0.0000304<br>(0.000) |
| Observations | 27448 | 30785 | 25052 | 30821 | 31203 |
| Temp. controls | Yes | Yes | Yes | Yes | Yes |
| Humidity controls | Yes | Yes | Yes | Yes | Yes |
| Precipitation controls | Yes | Yes | Yes | Yes | Yes |
| Wind speed controls | Yes | Yes | Yes | Yes | Yes |
| FE County X Year | Yes | Yes | Yes | Yes | Yes |
| FE Month | Yes | Yes | Yes | Yes | Yes |
| FE County X Inf. Seas. | No | No | No | No | No |

Notes: The dependent variable is the count of hospital admissions with diagnosed influenza within a county and month in Column (1). The outcomes in the following columns 2-5 are: Diabetes mellitus with complications, Urinary tract infections, Skull and face fractures and Osteoarthritis. We include only the influenza intensive months of October through March. Results are from a Poisson Pseudo-Maximum Likelihood regression with specified fixed effects and control variables. The number of included observations can vary across different outcomes due to fixed effects and varied counts in each county-month cell. Temperature controls consist of three separate bins, specific humidity controls consist of five separate bins, and precipitation and wind speed are linear terms. All weather variables are based on the monthly county averages. A higher AQI means worse air quality. Standard errors in parentheses are clustered at the county level.

**Table S.4:**
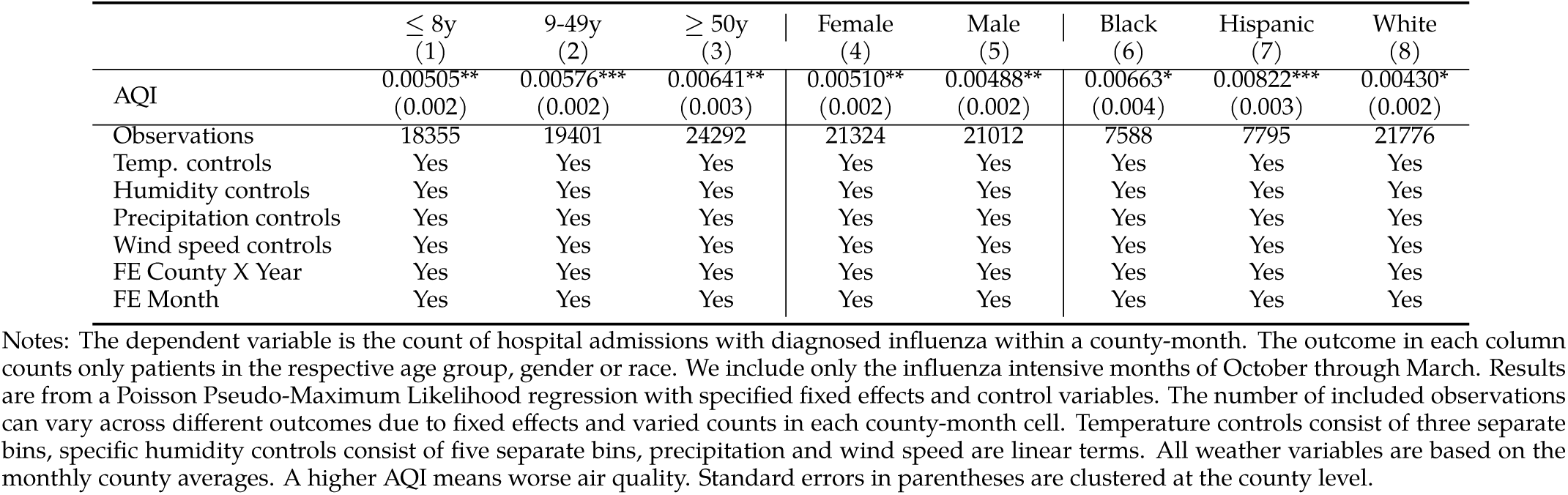
The effect of air pollution on influenza hospitalizations: heterogeneity

|  | ≤ 8y<br>(1) | 9-49y<br>(2) | ≥ 50y<br>(3) | Female<br>(4) | Male<br>(5) | Black<br>(6) | Hispanic<br>(7) | White<br>(8) |
| --- | --- | --- | --- | --- | --- | --- | --- | --- |
| AQI | 0.00505**<br>(0.002) | 0.00576***<br>(0.002) | 0.00641**<br>(0.003) | 0.00510**<br>(0.002) | 0.00488**<br>(0.002) | 0.00663*<br>(0.004) | 0.00822***<br>(0.003) | 0.00430*<br>(0.002) |
| Observations | 18355 | 19401 | 24292 | 21324 | 21012 | 7588 | 7795 | 21776 |
| Temp. controls | Yes | Yes | Yes | Yes | Yes | Yes | Yes | Yes |
| Humidity controls | Yes | Yes | Yes | Yes | Yes | Yes | Yes | Yes |
| Precipitation controls | Yes | Yes | Yes | Yes | Yes | Yes | Yes | Yes |
| Wind speed controls | Yes | Yes | Yes | Yes | Yes | Yes | Yes | Yes |
| FE County X Year | Yes | Yes | Yes | Yes | Yes | Yes | Yes | Yes |
| FE Month | Yes | Yes | Yes | Yes | Yes | Yes | Yes | Yes |

Notes: The dependent variable is the count of hospital admissions with diagnosed influenza within a county-month. The outcome in each column counts only patients in the respective age group, gender or race. We include only the influenza intensive months of October through March. Results are from a Poisson Pseudo-Maximum Likelihood regression with specified fixed effects and control variables. The number of included observations can vary across different outcomes due to fixed effects and varied counts in each county-month cell. Temperature controls consist of three separate bins, specific humidity controls consist of five separate bins, precipitation and wind speed are linear terms. All weather variables are based on the monthly county averages. A higher AQI means worse air quality. Standard errors in parentheses are clustered at the county level.

**Table S.5:**
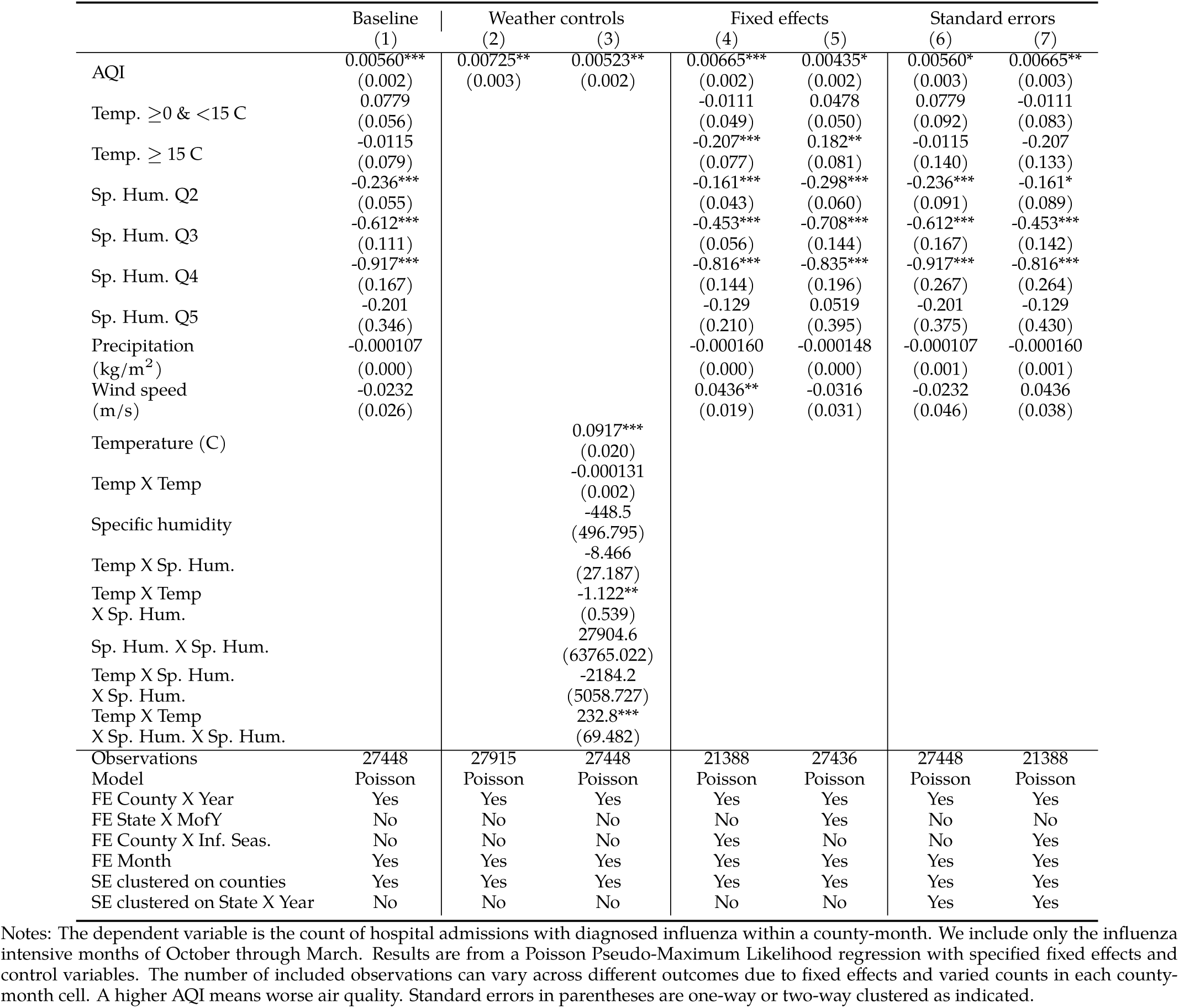
Robustness checks: different controls, fixed effects and standard errors

**Table S.6:**
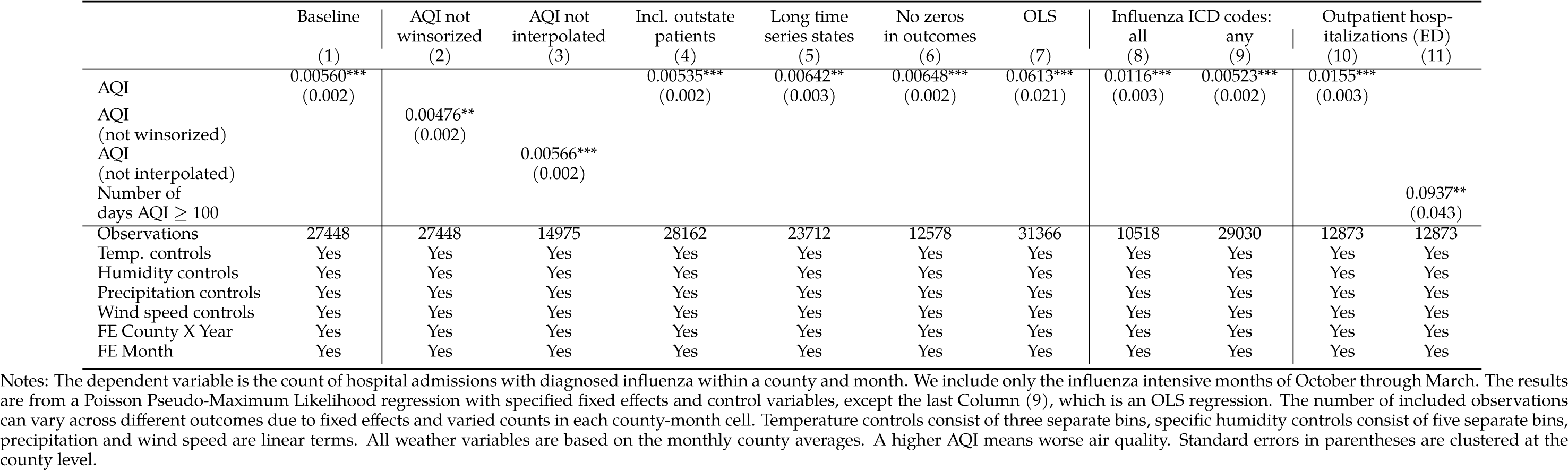
Further robustness checks

